# Human papillomavirus (HPV) prevalence in relation with cervical cytology in Bengali population of India

**DOI:** 10.1101/2020.06.01.20119628

**Authors:** Puja Banerjee, Arghya Bondhopadhyay, Bibek Mohan Rakshit, Amitava Pal, Anupam Basu

**Author notes:** Corresponding Author: **Dr. Anupam Basu, Professor**, Department of Zoology, The University of Burdwan, Golapbag, Burdwan-713104.

## Abstract

**Background:** Human papillomavirus (HPV) is one of the major infectious agents of cervical cancer. Papanicolaou (pap) smear study is generally carried out to screen the initial cervical condition and consequently specific PCR based study is carried out to recognize the different types of HPV. In the present study, we would like to screen the frequency of HPV infection in the women with normal and abnormal cervical discharges.

**Methods:** In our study, 216 subjects were recruited. Cervical cytology was done by Pap smear test. Nested PCR was carried out using MY09/11 and GP 5+/6+ primers to screen HPV infection.

**Result and conclusion:** A significant co-relation between HPV infection and early sexual intercourse was observed. We found a higher HPV prevalence in the age group below 29 years(35.48%). 85.71% SCC patients were positive for HPV infection, 80% HSIL patients were positive for HPV infection, 75% LSIL patients were positive for HPV infection; 66.7% ASCUS patients were positive for HPV infection. 50% ASC-H patients were positive for HPV infection. HPV positive was found in 22.22% of the subjects, among them 16.75% show normal cytology (NILM).

## Introduction

Cervical cancer is one of the major threats of death of women worldwide with human papillomavirus(HPV) recognized as its major causative agent (1). In India, approximately 1,34,420 women are diagnosed with cervical cancer every year, and 72,825 of them die due to the disease (2). HPV is a double stranded DNA virus which infects the squamous epithelium of the uterine cervix. More than 100 types are known to occur that are categorized into three major classes depending upon their oncogenic potential: high-risk types including HPV-16, 18, 31, 33, 35, 39, 45, 51, 52, 56, 58, 59, 68, 73 and 82; intermediate types are HPV-26, –53, –66 and low-risk types are HPV-6, –11, –40, –42, –43, –44, –54, –61, –70, –72, –81 and –CP6108 (3). Accordingly, Harald ZurHausen in the 1970s, HPV play a vital role in the development of cervical cancer, many observational studies (molecular, epidemiological and clinical) clearly implicating HPV as an etiological factor in a variety of anogenital cancers, as well as the cervix (4). The key attention in HPV relates to its contributory role in cervical cancer, one of the most common cancers in women, with an annual prevalence of approximately half a million and a mortality rate of approximately 50% worldwide (5).

The global distribution of cervical cancer extends, with Asia, Africa, Latin America exhibiting a higher rate of burden of this disease (6, 7). Around 11.4% women in the common population are estimated to have HPV infection (8). The highest frequency of HPV is located in sub-Saharan African regions followed by Latin American and Caribbean regions, Eastern Europe and South-eastern Asia. The incidence of particular HPV types in cervical malignancies may differ with the geographic basis of the specimen. In developing countries like India, occurrence of Human Papillomavirus (HPV) and their types was detected to be far above the ground. According to Globocan database throughout India, the predictable 5-year frequency of cancer in the adult female population for all cancers is 1,125,960, of which cervical cancer represents 308,901 (27.4%) (IARC 2012) (9). Studies on frequency of HPV and types related to cervical cancer have been conducted in different parts of the country. Cervical cancer is prevalent in north-eastern and eastern regions also. In the north-eastern states of India like Sikkim and Manipur a comparative profile of the prevalence of human papillomavirus type 16/18 infections in relation with age was done (10). In South 24-Pargana and West Bengal some researchers have found in case of cervical cancer that mostly HPV 16, 18, 33 are prevalent types (11).

High frequency of HPV42 and HPV56 has been observed in the southern side of the Tamil Nadu region (according to previous IARC surveys) (12). On the other hand, in Chennai, capital for the Tamil Nadu HPV16 was known as the most common type of high risk HPVs causing cervical lesions. (13).

Cervical cancer detection is initially done by Papanicolaou test. The Papanicolaou (Pap) test is a selection analysis conducted with cells from the uterine cervix. In 1943 George Papanicolaou initiated the Pap test as a cervical screening test. The test is easy, rapid, and trouble-free. The major utilization of orange G6 in the papanicolaou stain is to stain the keratin (14). Cervical cytology test is capable of detecting cervical cancer at premature phase and is broadly used in urban countries, where it has reduced both the frequency and death of cervical cancer. It has been evaluated that the utilization of this easy and cost-effective technique has minimized the occurrence of cervical cancer by at least 70%. (15,16,17).

Pre-cancer and cancer are determined by histopathological tests through microscopic study of the organization of the cells in biopsy or surgical tissue sections. These two techniques have contributed enormously in decreasing the load of cervical cancer. A well-built correlation between HPV and cervical cancer inspired the development of several diagnostic tests, mainly those based on molecular biology.

Many researchers have shown that HPV infection may correlate with pap positive or negative samples (18). On the contrary, there are also many investigators do not showed any relationship between HPV infection and cervical cytology (19,20).

Till now, there are no reports available on the study of the direct correlation between different stages of cervical cytology with HPV infection. So, the aim of our present study is to determine the correlation between cervical cytology with HPV infection in precanzcerous and cancerous cervix in studied population of India.

## MATERIALS AND METHODS

This work has been done in between 2012 to 2016. Cervical smear samples were collected from 216 women attending the O.P.D of Department of Gynecology and Obstetrics, Burdwan Medical College and Hospital, Burdwan, West Bengal by experience Gynecologists. The study was approved by the Institutional Ethical Committee, The University of Burdwan. The patients having complains of white discharge, post coital or post-menopausal bleeding and lower abdominal pain, without any history of previous treatment for cervical lesions were included in the study. Whereas the patients previously treated for cervical lesions and undergoing radiotherapy and pregnancy were excluded from the study. Cervical smears were obtained for HPV detection and cytological analysis in all the cases.

### Patient Information

All the patients were clustered in terms of age distribution, parity and age at first intercourse and studied for HPV infection and cervical cytology in the respective group.

### PAP smear collection

PAP smears were collected from the ectocervix and endocervix with a wooden spatula and cytobrush. Cervical smear was further collected to the tube containing PBS for DNA extraction and also on the slide for cytology.

### PAP smear test

Cervical cytology was done by PAP smear test. The PAP smear was fixed in 95% alcohol or 100% methanol. After fixation smears were dipped in haematoxylin (preheated 60°C) for nucleus staining and then washed with tap water and acetic acid. After washing, the slides were dipped in OG-6, EA-50 for counter staining for few seconds followed by washing with acetic acid, methanol and xylene. The slides were then mounted with DPX. Blotting was done after each step (21). All cervical smears were Papanicolaou stained and observed for koilocytosis, keratinization, binucleation, atypical cell morphology according to Bethesda system 2001.

### DNA extraction

PBS solution containing samples were vortexed and then transferred into 1.5 mL tubes for DNA extraction. DNA was extracted from each cervical scrape sample using standard phenol-chloroform procedures. The DNA containing tubes were stored at –20^0^C for PCR detection.

### Detection of HPV infection: PCR using MY09/11 and GP 5+/6+ primers

For the detection of HPV infection, nested PCR was done using MY09/11 and GP 5+/6+ primers(22). The first amplification reaction was performed using MY09/11 primer with commercial PCR kit as follows: Denaturation 94^0^C for 45 sec, annealing at 48^0^C for 45 sec and extension at 72^0^C for 1 min; Final reaction volume was of 25 μL and amplified for 30 cycles. Second amplification reaction of the Nested PCR was performed using the first PCR product as template using GP 5+/6+ primers. The amplification reaction was performed as follows: Denaturation 94^0^C for 45 sec, annealing at 47^0^C for 45 sec and extension at 72^0^C for 1 min; Final reaction volume was of 25 μL and amplified for 30 cycles. The amplification products were eluted on 2% agarose gel stained with ethidium bromide and visualized by GelDoc Imaging System (UVP).

### Statistics

Statistical analysis was done using Graphpad Prism (version 5.0) software. Correlations among the groups were calculated using contingency table χ^2^ test and P< 0.05 was considered as significant.

## RESULT

### Patients’ information

In the present study, some of the demographic factors like the age, parity, age at first intercourse etc of the subjects are studied. The age distribution of the 216 subjects studied has been divided into four groups: 29 years and below, 30–39 years, 40–49 years and 50 years and above with maximum number of patients in the age group between 30–39 years. The parity (number of child birth) of the studied subjects ranges from 1 to 4 and above. For the last demographic factor studied i.e., age at first intercourse, 5 groups were made: below 17 years, 17–20 years, 21–25 years, 26–30 years and 31–35 years. The demographic factors studied are presented in Table 1.

**Table 1:**
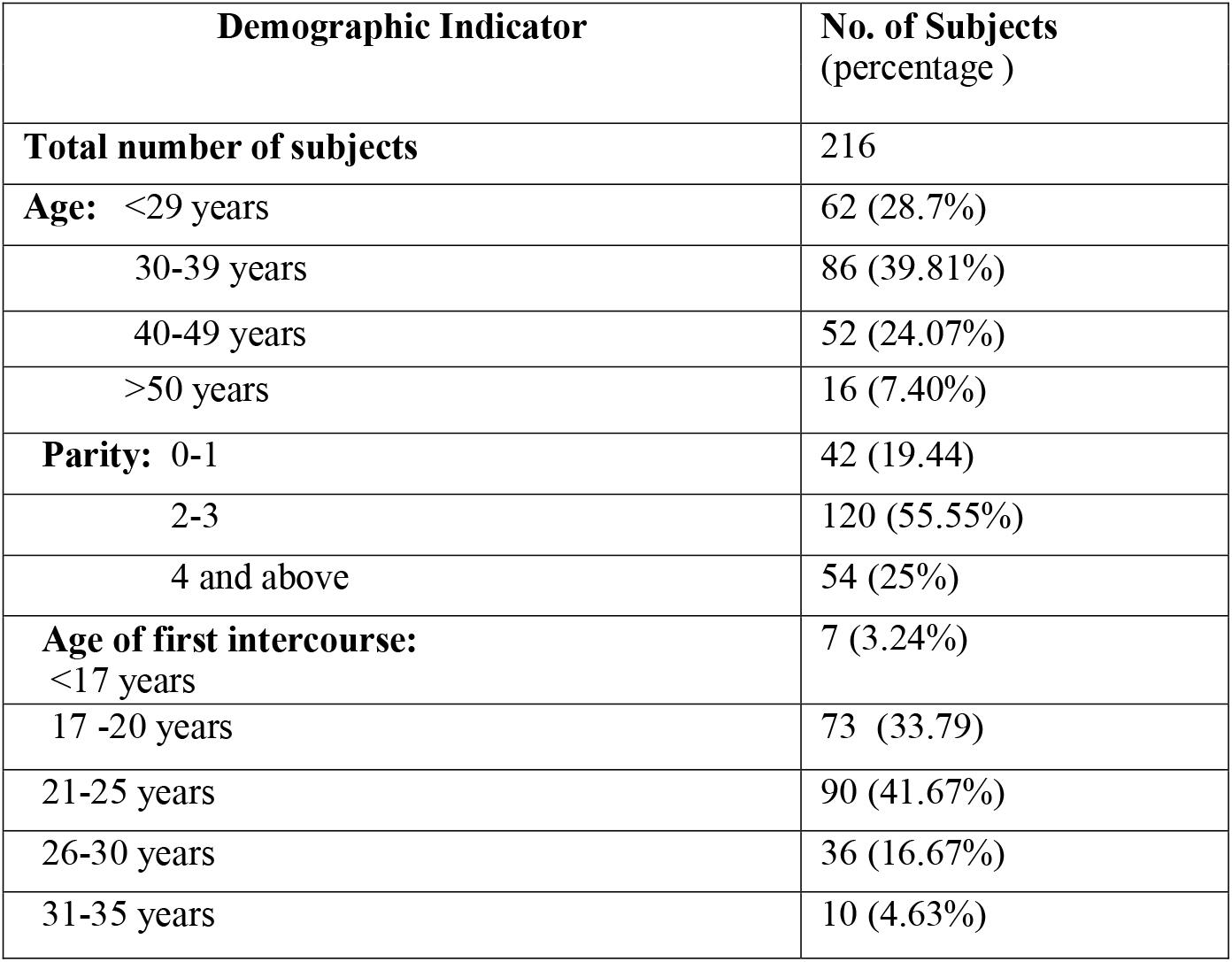
Different demographical information of the studied population.

| Demographic Indicator | No. of Subjects<br>(percentage ) |
| --- | --- |
| **Total number of subjects** | 216 |
| **Age:** <29 years | 62 (28.7%) |
| 30-39 years | 86 (39.81%) |
| 40-49 years | 52 (24.07%) |
| >50 years | 16 (7.40%) |
| **Parity:** 0-1 | 42 (19.44) |
| 2-3 | 120 (55.55%) |
| 4 and above | 54 (25%) |
| **Age of first intercourse:**<br><17 years | 7 (3.24%) |
| 17 -20 years | 73 (33.79) |
| 21-25 years | 90 (41.67%) |
| 26-30 years | 36 (16.67%) |
| 31-35 years | 10 (4.63%) |

### Cervical Cytology

PAP test was performed using OG-6 and EA-50 for the determination of cytological status of 216 subjects considered in the present study. As per Bethesda Classification, cervical cytology has been determined and presented in Fig1. Among 216 subjects 3.24% were positive for Squamous Cell carcinoma (SCC), 2.31% were positive for High Grade Squamous Intra-epithelial Lesion (HSIL) and 1.85% was positive for Low-Grade Squamous Intraepithelial Lesion (LSIL); 1.39% was positive for atypical squamous cells of undetermined significance (ASCUS) and 0.93% was positive for Atypical Squamous Cells, Cannot Rule out High-Grade Squamous Intra-epithelial Lesion (ASC-H). 16.75% subjects were normal cytology (NILM) (Table 2).

**Fig 1:**
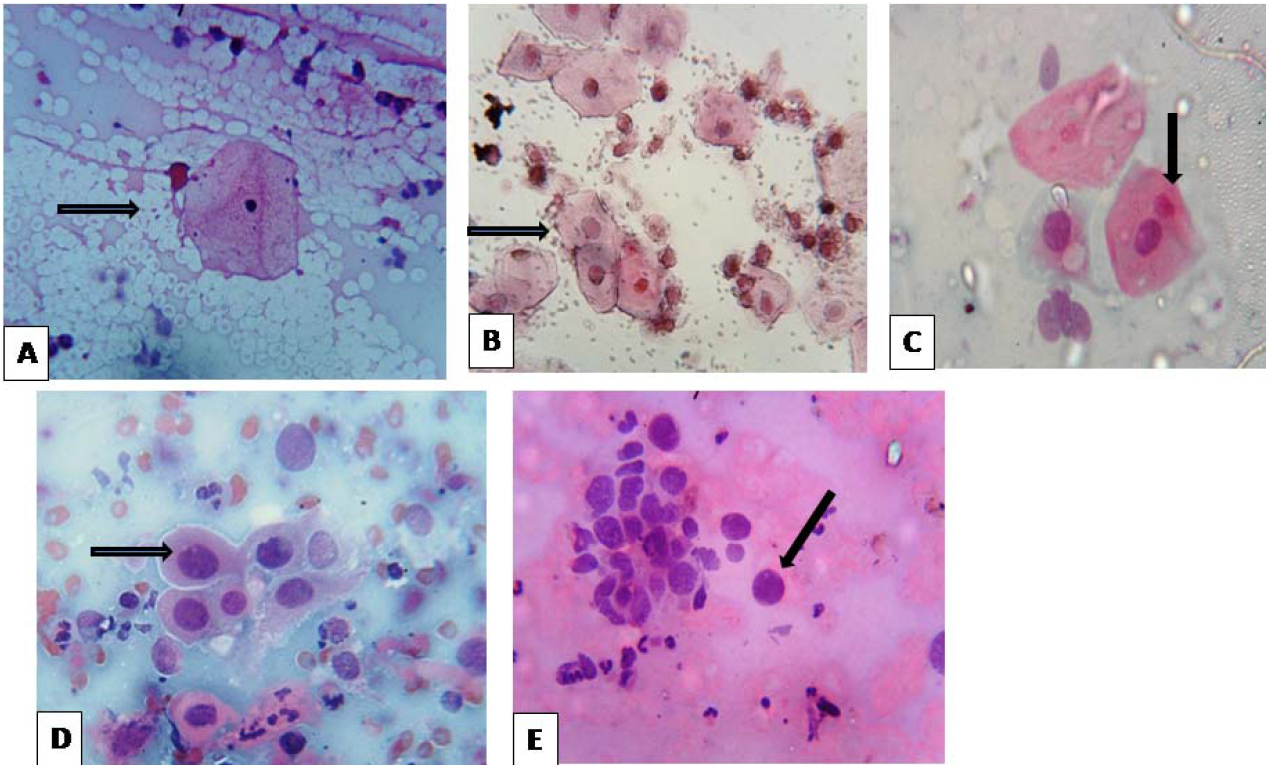
Cervical cytological findings as determined by the PAP stain using OG-6 and EA-50 **A:** Ectocervicalcytology from normal women of active reproductive age, showing the presence of the mature squamous epithelium cells. **B**: atypical squamous cells of undetermined significance (ASC-US) **C:** Low-Grade Squamous Intra-epithelial Lesion (LSIL); **D**: High-Grade Squamous Intra-epithelial Lesion (HSIL); **E:** Squamous Cell carcinoma (SCC).

**Table 2:** Cervical cytological findings in the studied population by PAP staining.

| Cytological finding* | No. of Subject |
| --- | --- |

|  | (Percentage) |
| --- | --- |
| NILM | 195 (90.27%) |
| ASC-H | 2 (0.93%) |
| ASC-US | 3 (1.39%) |
| LSIL | 4 (1.85%) |
| HSIL | 5 (2.31%) |
| SCC | 7 (3.24%) |

\* **NILM**- Negative for Intraepithelial Lesion or Malignancy, **ASC-H** -Atypical Squamous Cells, Cannot Rule Out High-Grade Squamous Intra- epithelial Lesion, **ASCUS** - Atypical squamous cells of undetermined significance, **LSIL** -Low-Grade Squamous Intra-epithelial Lesion, **HSIL** -High Grade Squamous Intra-epithelial Lesion, **SCC** -Squamous cell carcinoma

### HPV Infection and association with cervical cytology and different demographic relation

All 216 samples were tested for MY09/11 and GP5+/6+ primer sets by nested PCR (Fig.2). Among total of 216 studied samples, 48 were found HPV positive. In terms of correlation with cervical cytology with HPV infection, it has been observed that 85.71 % of SCC subjects were positive for HPV infection, 80% of HSIL were positive for HPV infection, 75 % LSIL subjects were found positive for HPV infection, 66.7 % of ASC-US subjects were positive for HPV infection and 50% of ASC-H subjects were found positive for HPV infection. Surprisingly it has been observed that approximately 16.75% of NILM cases showed HPV positive (Fig 3, Table 3). In the present study it has been observed that highest frequency of HPV infection was found in the age group below 29 years. The subjects with 2–3 parity show the higher chance of HPV infection. It has also been observed that subjects with early age of sexual activity, has higher chance of HPV infection. There was a significant association between age and HPV infection observed. There was also a significant association between HPV infection and early age of first intercourse observed. No significant association was found in case of parity in relation with HPV infection (Table 4).

**Fig 2:**
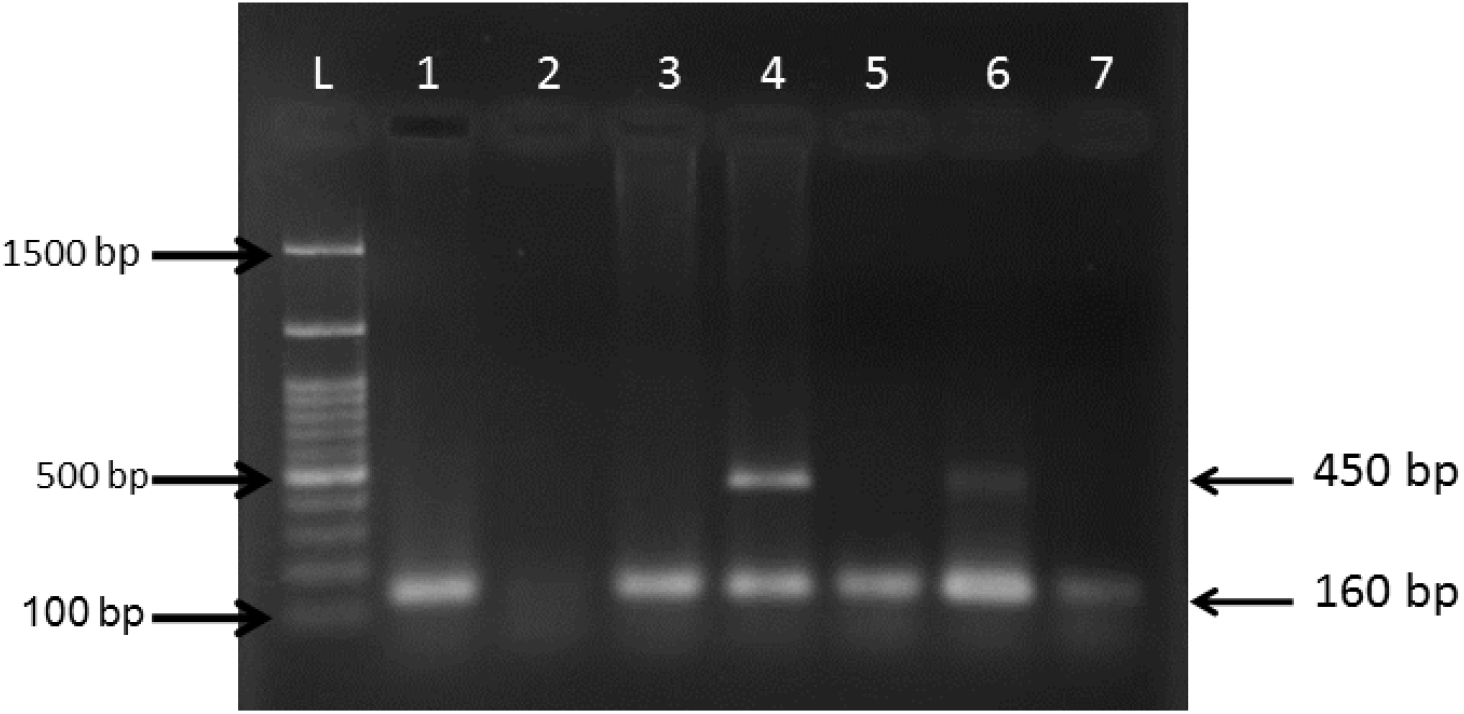
Polymerase chain reaction for the detection of HPV by nested PCR using MY09/11andGP5+/6+ primers resulting in amplicon size of 450bp and 160 bp respectively. (PCR products were eluted in 2% Agarose gel. Lane L :100bp ladder, Lane1,3,5,7:Ampplifed with MY09/11 followed by positive for GP5+/6+ Lane 2:Negative Sample, Lane 4,6: positive for My09/11 and GP5+/6+)

**Fig 3:**
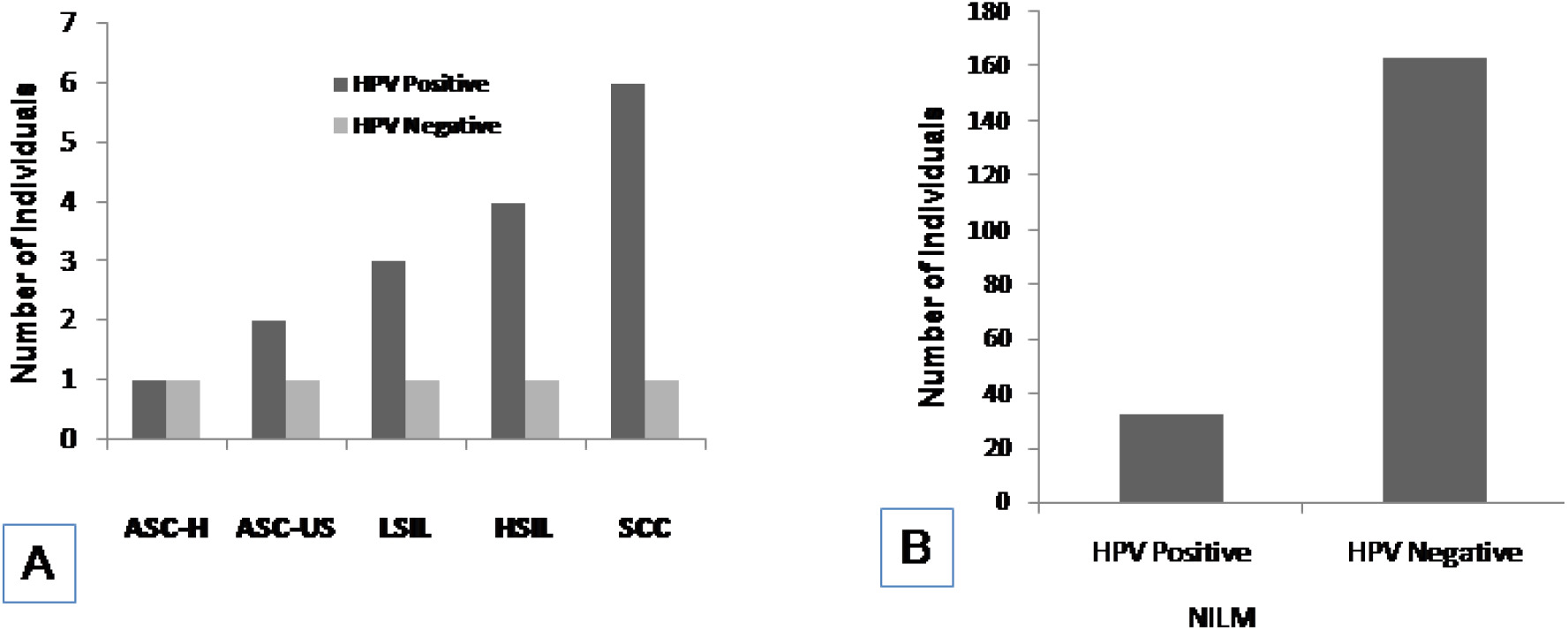
Cervical cytology in relation with HPV infection: A. HPV infection in relation with ASC-H, ASC-US, LSIL, HSIL and SCC subject; B. HPV infection in associated with NILM subjects

**Table 3:** Cervical cytology in relation with HPV infection in studied population.

| Cytological finding* | No. of Subjects | HPV Infection |  | Level of significance |
| --- | --- | --- | --- | --- |
|  |  | Positive frequency (%) | Negative frequency (%) |  |
| NILM | 195 (90.27%) | 32 (16.75%) | 163 (83.2%) | P<0.0001 |
| ASC-H | 2 (0.93%) | 1 (50%) | 1 (50%) |  |
| ASC-US | 3 (1.39%) | 2 (66.7 %) | 1 (33.3%) |  |
| LSIL | 4 (1.85%) | 3 (75%) | 1 (25%) |  |
| HSIL | 5 (2.31%) | 4 (80%) | 1 (20%) |  |
| SCC | 7 (3.24%) | 6 (85.71%) | 1 (14.28%) |  |

**Table 4:**
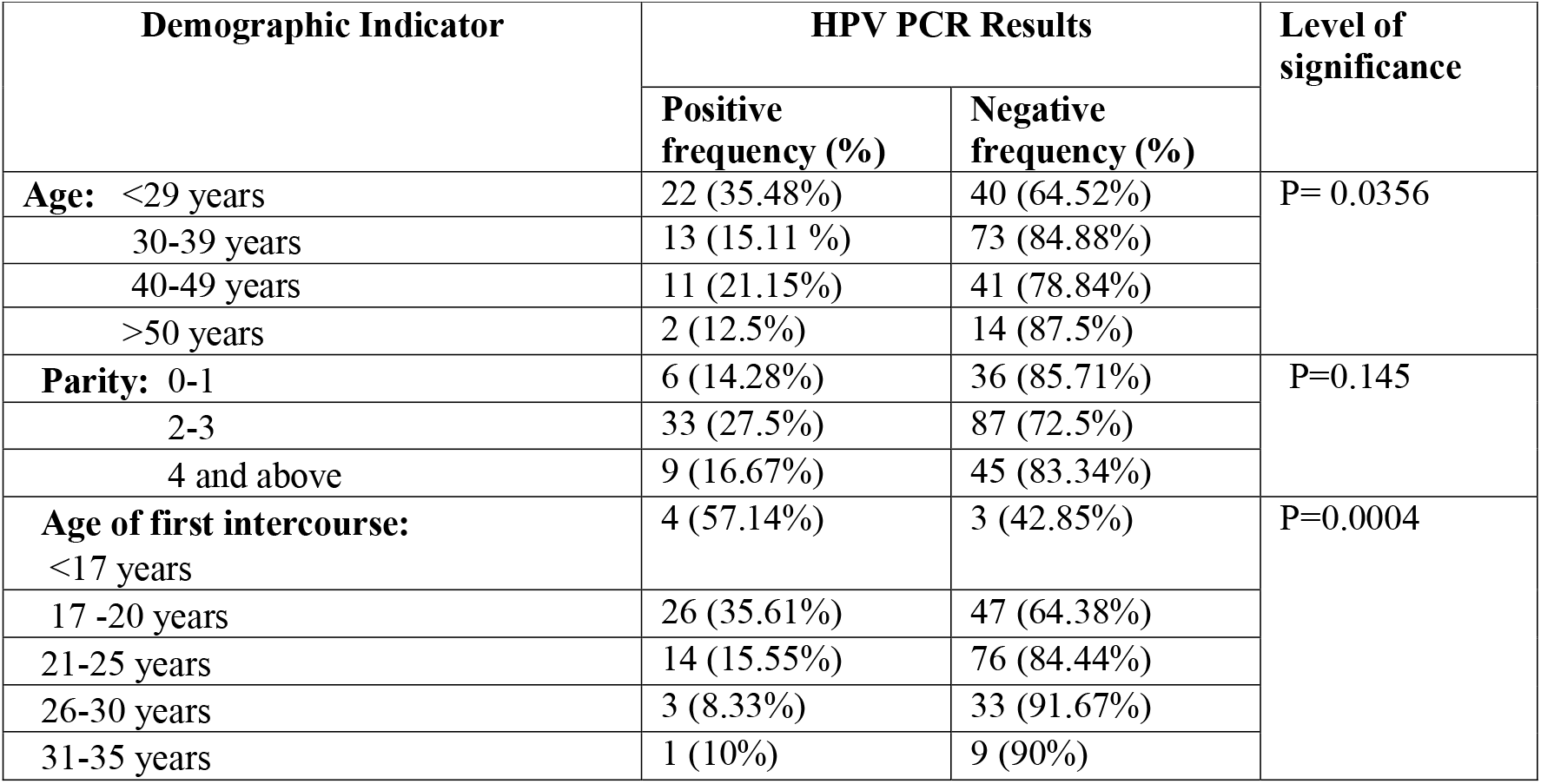
HPV infection in correlation with different demographic factors.

## DISCUSSION

In the present investigation, it has been studied that correlation between cervical cytology along with HPV infection. Cervical cancer is one of the major threats in women worldwide. Early detection of cervical cancer is possible through cytological screening and or HPV screening in regular interval. Several reports are available where it has been shown that HPV infection is prevalent up to age of 35 years in different populations (23–27). But age may not be the only criteria for defining HPV infection. Several factors are also responsible for the occurrence of HPV infection.

HPV is one of the major risk factor in the south eastern countries. Southeast Asian countries like, Indonesia, Philippines and Thailand have low frequency of HPV (9–16%) (28–30). In India, frequency of HPV varies in different populations. It has been reported that in Manipur, another state of eastern India, HPV prevalence is very low (7.4%). Thus in the present study we have investigated the prevalence of HPV and it’s correlation with PAP’s cytology in Bengali Indian population. Bengalis are mainly residing the West Bengal state of India. Different demographic indicators like – age of first intercourse and parity has also been studied for the HPV infection in this population group.

In the present investigation, it is found that higher HPV prevalence in the age group below 29 years (35.48%). Previously, it was also reported that predominance of HPV was found in younger age (23). We observed a significant association between HPV infection and first sexual intercourse at age group below 17 years. According to various studies it has been reported that early sexual intercourse was associated with HPV infection. Thus, our observation is well supported by the earlier report from different world populations (24–26).

Parity may influence HPV infection. Though previously, Srivastava *et al* 2014, showed that high parity (>3) is one of the important risk factors for HPV in the Indian population (27), but in the present study there is no significance association between parity and HPV infection has been observed. This may be due to the local habit, hygienic condition as well as socio-economic background.

Cytological examination of the cervix, i.e., PAP smear, is one of the major techniques in cervical cancer screening in most of the countries. In the present work, it has been observed that HPV infection is related to the different stages of cervical cytology. We have found that 85.71% of the squamous cell carcinoma subjects showed HPV infection. A significant association was observed between cervical cytology and HPV infection. Our observation in Bengali population is also correlated with other Indian population (27). The present findings of higher prevalence of HPV in the squamous cell carcinoma subjects also well supported by the findings from the population of Paraguay, Italy and Venezuela where 97%, 90.3% and 98.7% of the SCC patients showed HPV infection (31–33).

Surprisingly, in our study we have observed that 16.75% of the patients with normal cytology (NILM) have HPV infection. Our findings is well supported by the earlier studies by Yuan *et al*., 2011; Dondog *et al*., 2008; Keita *et al*., 2009, where it has been reported the infection of HPV with normal cervical cytology in Chinese, Mongolian and Guinea population respectively (34–36). These HPV positive women are prone to the development of cervical cancer and therefore should be examined carefully. Thus, routine screening is very important for the women for management of cervical cancer. This type of silent HPV infection, if it is remains unattended, may responsible for the future development of cervical cancer.

The present work, perhaps, may be the first time demonstrating the occurrence of HPV infection in women with normal cervical cytology, in Bengali, Indian population. Thus, the present study may be helpful in understanding the importance of regular cervical screening of the asymptomatic women for detecting of silent HPV infection in the uterine cervices, in the country like India with lower economic group people.

## Data Availability

as per request

## Acknowledgments

Authors express their thanks to DST- FIST program for providing different facilities to carry out the research

## Conflict of Interest

There is no conflict of interest to disclo

## Abbreviation

SCC: Squamous cell carcinoma
HSIL: High Grade Squamous Intra-epithelial Lesion
LSIL: Low-Grade Squamous Intra-epithelial Lesion
ASCUS: Atypical squamous cells of undetermined significance
ASC-H: Atypical Squamous Cells, Cannot Rule Out High-Grade Squamous Intra-epithelial Lesion
NILM: Negative for Intraepithelial Lesion or Malignancy

## Notes

### Competing Interest Statement

The authors have declared no competing interest.

### Funding Statement

in house

### Author Declarations

institutional ethics committee, Burdwan University

